# Breath volatile profiling reveals a diagnostic signature of MASLD in children

**DOI:** 10.64898/2026.05.26.26353794

**Authors:** Amalia Z. Berna, Jennifer Panganiban, Yang Liu, Joey Logan, Pierre Russo, Aakanshya Aryal, Kathryn Hafertepe, Sarah Abu-Alreesh, Brian DeBosch, Janis Stoll, Audrey R. Odom John

**Author notes:** Corresponding author: Audrey R. Odom John. these authors contributed equally (Amalia Z. Berna, Jennifer Panganiban).

## Abstract

**Background & Aims:** Metabolic Dysfunction–Associated Steatotic Liver Disease (MASLD) is the leading cause of chronic liver disease in children. However, accurate, noninvasive diagnostic tools remain limited. Current screening methods are invasive or lack sensitivity. Breath-based volatile organic compound (VOC) analysis offers a simple approach with potential for point-of-care screening. This study aimed to identify and validate breath VOC signatures of pediatric MASLD.

**Approach & Results:** We conducted a prospective IRB-approved cohort study at the Children’s Hospital of Philadelphia (CHOP). Children aged 7–20 years with MASLD (n=22), as defined by hepatic steatosis either by liver biopsy or imaging and 1 cardiometabolic risk factor, and a control group without MASLD (n=20) were enrolled. Breath samples were collected using a standardized protocol and analyzed by untargeted comprehensive two-dimensional gas chromatography-mass spectrometry (GC×GC-MS). Machine learning and unsupervised clustering were applied to identify discriminatory VOCs and assess heterogeneity. Untargeted GC×GC-MS analysis identified a distinct breath VOC signature in children with MASLD compared with non-MASLD controls. A Random Forest model achieved a sensitivity of 73% and specificity of 65%, with AUC of 0.84. The VOC 2,4-dimethyl-1-heptene demonstrated strong diagnostic performance in the discovery cohort with a sensitivity of 85%, specificity of 77% and an AUC of 0.81. Unsupervised clustering revealed four MASLD subgroups with distinct volatile phenotypes associated with differences in liver enzymes and metabolic parameters. External validation in a second pediatric cohort confirmed reproducible reductions in o/p-xylene in subjects with MASLD.

**Conclusions:** Pediatric MASLD is associated with a reproducible breath VOC signature identified by untargeted GC×GC-MS. These findings support breath analysis as a scalable, noninvasive screening and stratification tool for pediatric MASLD and warrant validation in larger, longitudinal studies.

**IMPACT AND IMPLICATIONS:** Pediatric MASLD lacks scalable, noninvasive diagnostic tools capable of early identification of at-risk children, providing strong scientific justification for the development of breath-based biomarkers. This study demonstrates that untargeted breath VOC profiling can distinguish children with MASLD from controls and reveals biologically meaningful heterogeneity in breath signatures, highlighting the potential of breath analysis for both detection and risk stratification. These findings are most relevant to physicians managing children with cardiometabolic risk and to researchers developing noninvasive liver biomarkers. Broader validation is required before clinical implementation.

## INTRODUCTION

Metabolic Dysfunction-Associated Steatotic Liver Disease (MASLD), previously called non-alcoholic fatty liver disease (NAFLD), is the leading cause of chronic liver disease in children. Using historic diagnostic criteria, MASLD has an estimated prevalence of 10-16% in children in the United States and a global prevalence of 5%. Higher rates are seen in males, older children, Hispanic or Asian ethnicity, and in children with obesity ^1^. Up to 17% of children with MASLD may have advanced fibrosis on diagnosis, with ∼23% experiencing fibrosis progression. Metabolic dysfunction-association steatohepatitis (MASH) has become the primary reason for liver transplant listing in young adults 18-40 years old, with significant increase in mortality in youth-onset MASLD.

Despite the increasing prevalence and risk for progression, pediatric MASLD remains underdiagnosed ^2^. The 2023 American Academy of Pediatrics (AAP) guidelines recommend MASLD screening in higher risk children ≥ 10 years of age, using sex-specific normal values for ALT (>22 IU/dL for girls and > 26 IU/L for boys). Despite this, only 27% of children with obesity receive AAP-adherent screening for MASLD, diabetes, and hyperlipidemia ^3^.

Based on the new nomenclature, elevated ALT alone is insufficient to diagnose MASLD, which requires confirmation of steatosis by imaging or liver biopsy and the presence of at least one cardiometabolic risk factor. Unlike in adults, there are no validated thresholds using serologic biomarkers or imaging noninvasive tests (NITs) for accurate detection of MASH and fibrosis in children. ^1^. Although magnetic resonance elastography (MRE) with proton-dense fat fraction (PDFF) is considered the most reliable imaging technique for quantitatively measuring steatosis, this is greatly limited by accessibility, cost, and possible inability to stay still in an MRI machine.

Human exhaled breath contains hundreds of volatile organic compounds (VOCs), including hydrocarbons, furans and ethers, nitrogen-containing compounds, sulfur compounds, ketones, and other oxygenated compounds ^4^. Pathological conditions can alter breath composition, which can be measured using precise analytical instruments like gas chromatography mass spectrometry (GC-MS). Analysis of exhaled breath VOCs holds potential as a noninvasive biomarker for liver disease diagnosis.

While prior studies have explored breath VOC changes in adults with various liver conditions ^5–9^, analogous studies in children are limited. One pediatric study by Alkhouri *et al* ^10^ demonstrated that VOC analysis using selective ion flow tube mass spectrometry (SIFT-MS) differentiated overweight or obese children with Non-alcoholic Fatty Liver Disease (NAFLD), diagnosed via ultrasound, from those without NAFLD. Among 14 preselected VOCs, five—isoprene, acetone, trimethylamine, acetaldehyde, and pentane—were significantly elevated in the NAFLD group. In a follow up study, the same research group ^11^ used SIFT-MS to compare the breath profiles of children with various chronic liver diseases (CLD) to healthy controls. Of the 21 preselected VOCs analyzed, seven showed significant differences between groups. Notably, breath levels of 1-decene, 1-heptene, 1-octene, and 3-methylhexane were significantly increased in children with advanced fibrosis relative to healthy participants. The use of targeted approaches (preselected VOCs) may overlook key diagnostic signals, especially in a disease as metabolically heterogeneous as MASLD. To address this limitation, we employ an untargeted two-dimensional gas chromatography-mass spectrometry (GC×GC-MS) approach to comprehensively characterize the pediatric breath volatilome. This method allows for a broader assessment of volatile organic compound (VOC) patterns associated with disease, with reduced bias.

We sought to identify breath-based biomarkers that may facilitate early, noninvasive detection and risk stratification of MASLD in children. Notably, all participants in our study met criteria for overweight/obesity and were classified as either non MASLD or MASLD based on liver ultrasound or confirmatory liver biopsy, ensuring well-defined clinical phenotypes.

## METHODS

### Study Approval and Enrollment

This study was designed as a prospective cross-sectional study at the Children’s Hospital of Philadelphia (CHOP). Prior to enrollment, the study was approved by the CHOP Human Research Ethics Committee (IRB 21-019125). Participants or their legal representatives consented to take part in the present study. Assent was obtained from children aged 7 years and older, in accordance with institutional guidelines.

Children with MASLD (aged 7–20 years) were enrolled from the CHOP MASLD Clinic. MASLD group inclusion criteria were as follows: (1) BMI ≥85% for age/sex based on CDC growth charts, hepatic steatosis on imaging and an alanine aminotransferase (ALT) ≥2x the upper limit of normal ALT for sex (≥52 U/L for males ≥44 U/L for females); or (2) biopsy-confirmed MASLD within 6 months of subject enrollment.

An age-matched “non MASLD” group, recruited from our Healthy Weight (Obesity) outpatient clinic, was also included in the study. Controls consisted of subjects who had BMI ≥85% for age/sex based on CDC growth charts, absence of hepatic steatosis on ultrasound, and normal liver enzymes (ALT ≤ 26 U/L for males; ≤ 22 U/L for females). Ultrasound studies were performed by a trained professional in the CHOP Radiology department.

Children were excluded from study if they had uncontrolled asthma or asthma requiring inhaler use within 48h prior, as well as those with neurological disorders, altered mental status, or who were otherwise unable to cooperate with voluntary collection. In addition, children using medications that could cause hepatic steatosis were excluded.

The results from this work were further evaluated with samples derived from an independently collected cohort at St. Louis Children’s Hospital (SLCH), where children under evaluation for MASLD and age-matched non MASLD subjects were enrolled. Prior to enrollment, the study was approved by SLCH Human Research Ethics Committee (IRB 201709030). In this cohort, all patients had a BMI ≥85% for age/sex based on CDC growth charts, and ALT ≥2× the upper limit of normal for sex in the suspected MASLD group and normal ALT in the non MASLD group.

### Histological Features

Biopsies were evaluated by a single pathologist who was blinded to clinical and laboratory data. Liver biopsies were graded according to NAFLD activity score (NAS) proposed by Kleiner et al ^12^ and fibrosis. NAS (scores 0-8) method combines the grade of steatosis (0-3), lobular inflammation (0-3), and ballooning (0-2). Fibrosis score was assessed 0-4. Stage 0 (no fibrosis); Stage 1a (zone 3 perisinusoidal fibrosis delicate); Stage 1b (zone 3 perisinusoidal fibrosis dense); Stage 1c (portal-only fibrosis, without perisinusoidal fibrosis); Stage 2 (as above with portal fibrosis); Stage 3 (as above with bridging fibrosis); and Stage 4 (cirrhosis).

### Clinical Parameters

Clinical metadata was extracted from the Electronic Medical Record. Data collected included demographic information (age, gender, race/ethnicity), anthropometrics (weight, height, BMI, BMI z-score, and BMI percentile) and related laboratory data, including the following: alanine aminotransferase (ALT), aspartate aminotransferase (AST), glucose (GLU), total bilirubin (T-Bil), alkaline phosphatase (ALP), gamma-glutamyltransferase (GGT) and total protein (TP) (**Table 1**). Data were collected within the last 9 months prior to sample collection.

**Table 1:** Characterization of study population by MASLD status (CHOP cohort)

| Variable | MASLD<br>N = 22<sup>1</sup> | Non MASLD<br>N = 20<sup>1</sup> | p-value<sup>2</sup> |
| --- | --- | --- | --- |
| **Clinical features** |  |  |  |
| Age | 15 (12.0, 17.0) | 13 (10.0, 14.5) | 0.084 |
| Sex |  |  | 0.006 |
| Female | 6 (27%) | 14 (70%) |  |
| Male | 16 (73%) | 6 (30%) |  |
| Race |  |  | <0.001 |
| Asian | 2 (9.1%) | 0 (0%) |  |
| Black or African American | 0 (0%) | 13 (65%) |  |
| Other | 11 (50%) | 1 (5.0%) |  |
| White American/ Caucasian | 9 (41%) | 6 (30%) |  |
| Ethnicity |  |  | 0.005 |
| Hispanic or Latinx | 11 (50%) | 2 (10%) |  |
| Not Hispanic or Latinx | 11 (50%) | 18 (90%) |  |
| BMI z-score | 2.33 (1.94, 2.51) | 3.02 (2.36, 3.32) | 0.016 |
| BMI | 34.0 (29.0, 39.2) | 36.6 (31.2, 41.0) | 0.3 |
| BMI percentile |  |  | 0.054 |
| 85<sup>th</sup> - 95<sup>th</sup> percentile | 4 (18%) | 3 (15%) |  |
| >95<sup>th</sup> percentile<br>n available | 18 (82%)<br>22/22 | 16 (80%)<br>19/20 |  |
| Glucose (mg/dL)<br>n available | 100 (98, 108)<br>16/22 | 89 (80, 92)<br>16/20 | <0.001 |
| ALT (U/L)<br>n available | 140 (96, 212)<br>22/22 | 16 (13, 18)<br>20/20 | <0.001 |
| AST (U/L)<br>n available | 79 (51, 97)<br>22/22 | 22 (16, 23)<br>20/20 | <0.001 |
| Total bilirubin (mg/dL)<br>n available | 0.50 (0.40, 0.70)<br>21/22 | 0.40 (0.20, 0.50)<br>15/20 | 0.029 |
| ALP (U/L)<br>n available | 177 (91, 252)<br>21/22 | 157 (111, 237)<br>15/20 | 0.9 |
| Total Protein (g/dL)<br>n available | 7.90 (7.60, 8.30)<br>21/22 | 7.30 (6.90, 7.70)<br>15/20 | <0.001 |
| GGT (U/L)<br>n available | 45 (37, 71)<br>2/22 | 13<br>1/19 | † |

<sup>1</sup>Median (Q1, Q3); n (%). <sup>2</sup>Wilcoxon rank sum test; Pearson's Chi-squared test; Fisher's exact test. ALT= Alanine aminotransferase. AST= Aspartate aminotransferase. ALP = Alkaline phosphatase. GGT = gamma-glutamyltransferase. † Statistical testing was not performed due to insufficient sample size in non MASLD group.

### Breath Collection

For both cohorts (CHOP and SLCH), breath samples were collected following previously described protocols ^13^. In brief, participants exhaled through a disposable cardboard mouthpiece connected to a collection chamber, which was linked via tubing to a 3-liter SamplePro FlexFilm sample bag (SKC Inc., Pennsylvania). Subjects were instructed to take several deep breaths, position the cardboard tube between their lips, and exhale fully into the chamber. No nose clips or VOC filters were used during the procedure. Breath samples were then transferred from the collection bags to sorbent tubes, following established methods ^14^. We used three-bed universal sorbent tubes containing Tenax, Carbograph, and Carboxen (Markes International Limited, UK). For each subject, a paired ambient air sample was collected from the same room and time point as the breath sample. All samples were stored at 4°C and analyzed within 2-4 weeks of collection. VOC analysis was performed via thermal desorption followed by two-dimensional gas chromatography and time-of-flight mass spectrometry (GC×GC BenchTOF-MS; SepSolve Analytical, UK), as described below.

### Breath Metabolite Analysis

Prior to analysis, sorbent tubes were equilibrated to room temperature and loaded into an autosampler (Ultra-xr, Markes International, UK). A gaseous internal standard mixture—consisting of 1.01 ppm bromochloromethane, 1.04 ppm 1,4-difluorobenzene, 1.04 ppm chlorobenzene-D5, and 0.96 ppm 4-bromofluorobenzene—was added to each tube. This was followed by a pre-desorption purge step using helium at 50 mL/min for 10 minutes to remove water content from the breath samples. Thermal desorption was carried out using a Unity-xr thermal desorber (Markes International, UK) at 270°C for 10 minutes. VOCs were then focused onto a “Universal” cold trap (sorbent-matched to the sample tube), initially held at 10°C and rapidly heated to 300°C to minimize peak broadening. The split flow after the cold trap was 3 mL/min.

Samples collected at CHOP were run using a two-dimensional gas chromatography (GC×GC) (Agilent 7890B GC system) equipped with a flow modulator (INSIGHT, SepSolve Analytical, UK) and a three-way splitter directing effluent to a flame ionization detector (FID) and a time-of-flight mass spectrometer (BenchTOF, SepSolve Analytical, UK). The first-dimension column was a Stabilwax (30 m × 250 μm i.d. × 0.25 μm df), and the second-dimension column was an Rtx-200MS (5 m × 250 μm i.d. × 0.1 μm df), both from Restek (Bellefonte, PA, USA). The oven temperature program was as follows: an initial hold at 40°C, ramped to 215°C at 3°C/min and held for 1 minute, followed by a rapid increase to 260°C at 50°C/min, with a final hold at 260°C for 10 minutes. The total run time was 70 minutes. Helium was used as the carrier gas at a constant flow of 1.2 mL/min. The flow modulator loop had dimensions of 0.53 mm i.d. × 110 mm (loop volume: 25 µL), with a modulation period of 2 seconds.

Breath samples collected from SLCH were run at Washington University in St Louis-SLCH on a GCqTOF (Agilent Technologies, USA) coupled to a Unity-xr thermal desorber (Markes International, UK) as previously described ^15^. Briefly, prior to analysis, sorbent tubes were equilibrated to room temperature, spiked with a gaseous standard mixture (bromochloromethane, 1,4-difluorobenzene, chlorobenzene-D5), and subjected to a 5-minute dry purge with nitrogen at 50 mL/min. Tubes were thermally desorbed at 270°C and analytes were recollected on a Tenax cold trap held at 10°C, then released at 295°C, with 20% of the sample transferred to a 7890B GC system (Agilent Technologies) equipped with a DB-5MS column (30 m × 0.25 mm × 0.25 µm). The oven temperature was held at 35°C for 3 minutes, increased to 175°C at 5°C/min, then ramped to 300°C at 60°C/min with a final 2-minute hold. Data were acquired at 50 Hz over a mass range of 35–400 m/z.

### Chemicals and Quality control

Pure standards of selected VOCs (**Table 2**) were purchased from Sigma-Aldrich and analyzed using the same GC×GC-TOF-MS protocol applied to breath samples. Quality control was performed by analyzing an external standard (EPA 8240B Calibration Mix) prior to each batch to monitor instrument stability over time, as previously described ^16^.

**Table 2:**
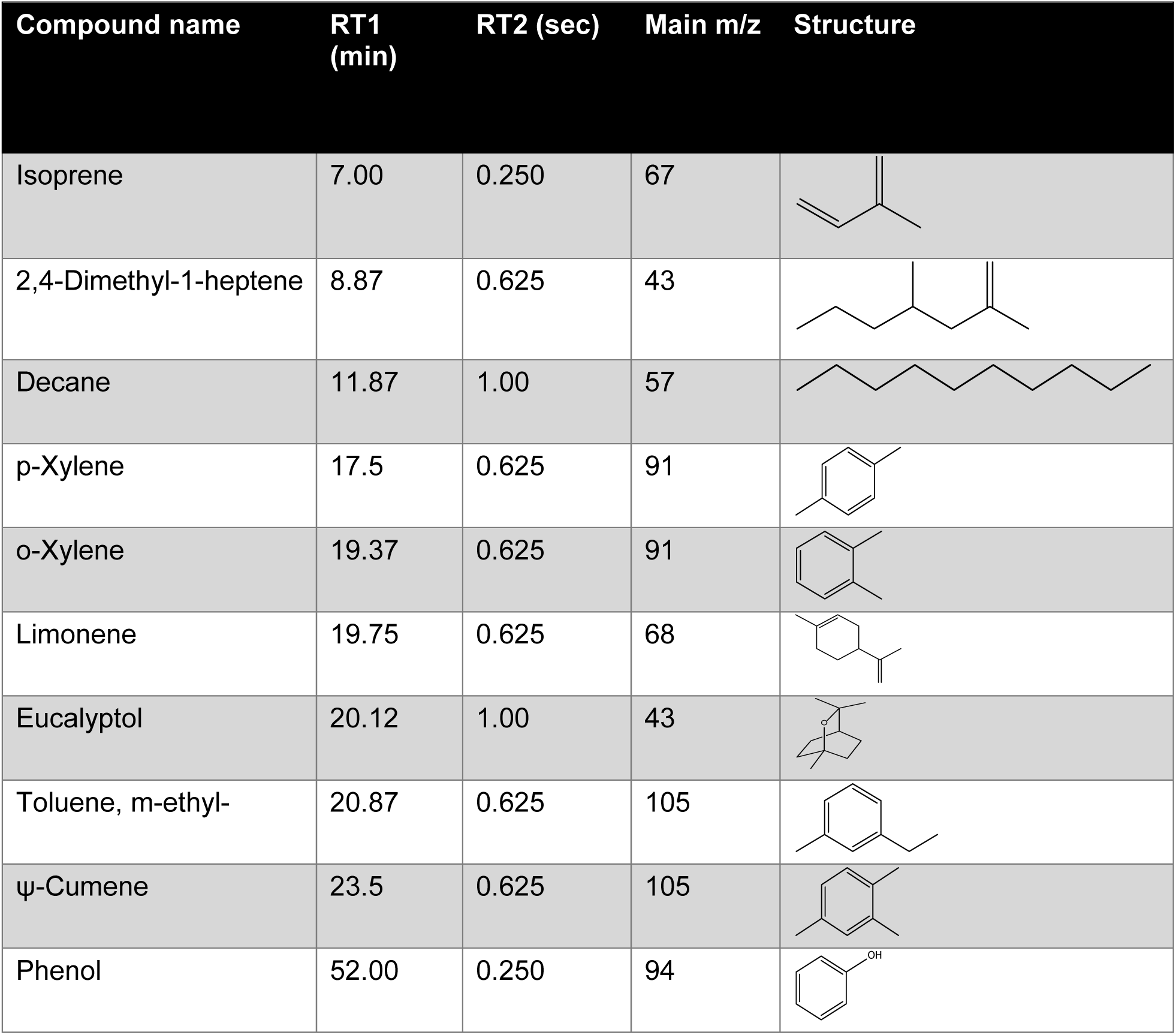
List of volatile organic compounds found to be discriminatory between MASLD and Non MASLD.

| Compound name | RT1 (min) | RT2 (sec) | Main m/z | Structure |
| --- | --- | --- | --- | --- |
| Isoprene | 7.00 | 0.250 | 67 |  |
| 2,4-Dimethyl-1-heptene | 8.87 | 0.625 | 43 |  |
| Decane | 11.87 | 1.00 | 57 |  |
| p-Xylene | 17.5 | 0.625 | 91 |  |
| o-Xylene | 19.37 | 0.625 | 91 |  |
| Limonene | 19.75 | 0.625 | 68 |  |
| Eucalyptol | 20.12 | 1.00 | 43 |  |
| Toluene, m-ethyl- | 20.87 | 0.625 | 105 |  |
| ψ-Cumene | 23.5 | 0.625 | 105 |  |
| Phenol | 52.00 | 0.250 | 94 |  |

### Untargeted Data Analysis

Raw GC×GC–MS chromatograms were aligned to a reference chromatogram using ChromCompare+ software (v2.1.4, SepSolve Analytical Ltd, UK). Following alignment, a tile-based approach was applied to extract features directly from the chromatograms without requiring prior peak integration or compound identification. Tiles were defined using a 32-second window in the first dimension and a 0.5-second window in the second dimension, with 25% overlap. For each tile, signal intensities for individual m/z channels were summed and compiled across all samples, resulting in a dataset of features indexed by retention time and m/z.

Features with m/z intensities below 30,000 and/or those suspected to be siloxanes or sorbent-related compounds were removed. Features were retained if they were present in over 50% of samples of at least one group. The resulting data were normalized to the internal standard (4-bromofluorobenzene) prior to analysis. Feature selection was conducted using the ChromCompare+ proprietary multivariate algorithm, which accounts for covariance among variables ^17^. This process reduced the dataset to the 50 most discriminative features distinguishing the groups under study. Among these, 10 features corresponded to unique volatile compounds. Preliminary identification of selected peaks was achieved through spectral matching against the NIST20 library, using a similarity threshold of ≥60%. Compound identities were confirmed by comparison to commercial analytical standards.

### Machine learning

In this study, we built a predictive model using a Random Forest classifier, an ensemble-based algorithm that generates multiple decision trees and combines their outputs to enhance prediction accuracy.

To evaluate model performance, we employed 5-fold cross-validation. The dataset was split into five equal parts, with each fold used once as a validation set while the remaining folds were used for training. This ensured that all data points were used for both training and testing, providing a more robust and unbiased estimate of model performance. Evaluation metrics—including accuracy, precision, recall, and area under the curve (AUC)—were averaged across the folds to assess overall predictive capability.

We further assessed feature importance using the Gini index, which allowed us to rank features by their contribution to reducing classification error across the trees.

### Statistical analysis

Unsupervised hierarchical cluster (HC) analysis (Euclidean distance, Ward’s linkage) was performed using discriminant MASLD biomarkers for all MASLD participants. Clusters generated were used to identify potential MASLD sub-groupings and to describe them based on breath concentrations and clinically relevant liver function variables. Cut points in the tree diagrams were chosen to avoid having clusters of fewer than 4 clusters, and to aim to have ≥8 subjects in at least two clusters. Spearman’s correlation analysis was applied to assess associations between VOCs and laboratory parameters and NAS or Fibrosis.

The discriminative performance of VOCs identified during untargeted analysis, following feature selection, was evaluated using receiver operating characteristic (ROC) curves.

Group comparisons of VOC levels were conducted using the nonparametric Mann-Whitney U test and box plots depict the median, with whiskers indicating the minimum and maximum values. Data visualization and statistical analysis, including cluster analysis, heatmaps, box plots, PCA and ROC curves, were carried out in RStudio (version 2024.04.1; Posit PBC, Boston, MA) and GraphPad Prism (version 10.4.0; GraphPad Software, San Diego, CA).

## RESULTS

A total of 42 patients were recruited from the Children’s Hospital of Philadelphia (CHOP) (**Table 1**). MASLD was diagnosed in 22 subjects based on the new MASLD nomenclature of hepatic steatosis on imaging or liver biopsy with at least 1 cardiometabolic risk factor. Twenty participants were in the non MASLD group. There was no significant difference in age between groups. Sex distribution differed between groups, as MASLD patients were predominantly male (73%), whereas non MASLD were predominantly female (70%) (p=0.006). Non MASLD subjects reported more Black or African American children (65%), compared to MASLD cases, and 50% of MASLD cases were Hispanic/Latinx versus 10% of non MASLD (p=0.005), with the remainder being non-Hispanic.

Median BMI z-score differed significantly between groups (p=0.016), with MASLD cases having a lower median (2.33) than non MASLD (3.02). Serum glucose levels (non-fasting) were higher in MASLD cases (median 100 mg/dL vs 89 mg/dL in controls; p<0.001). Liver transaminases were markedly elevated in MASLD and normal in non MASLD subjects, as expected as per inclusion criteria.

### Ten Volatile Compounds Discriminate MASLD Patients from Non MASLD

Two-dimensional GCxGC-MS analysis was used to investigate the breath profiles of subjects with and without MASLD. Using a feature selection algorithm, 10 VOCs were identified as discriminant markers of MASLD (**Table 2**). Of these VOCs, five VOCs were aromatic hydrocarbons. The levels of MASLD biomarkers were significantly elevated in non MASLD compared to MASLD subjects, except for isoprene (**Figure 1A**). Principal component analysis (PCA) used for visualization purposes revealed clusters of MASLD and control samples, indicating differences in their VOC profiles (**Figure 1B**).

**Figure 1:**
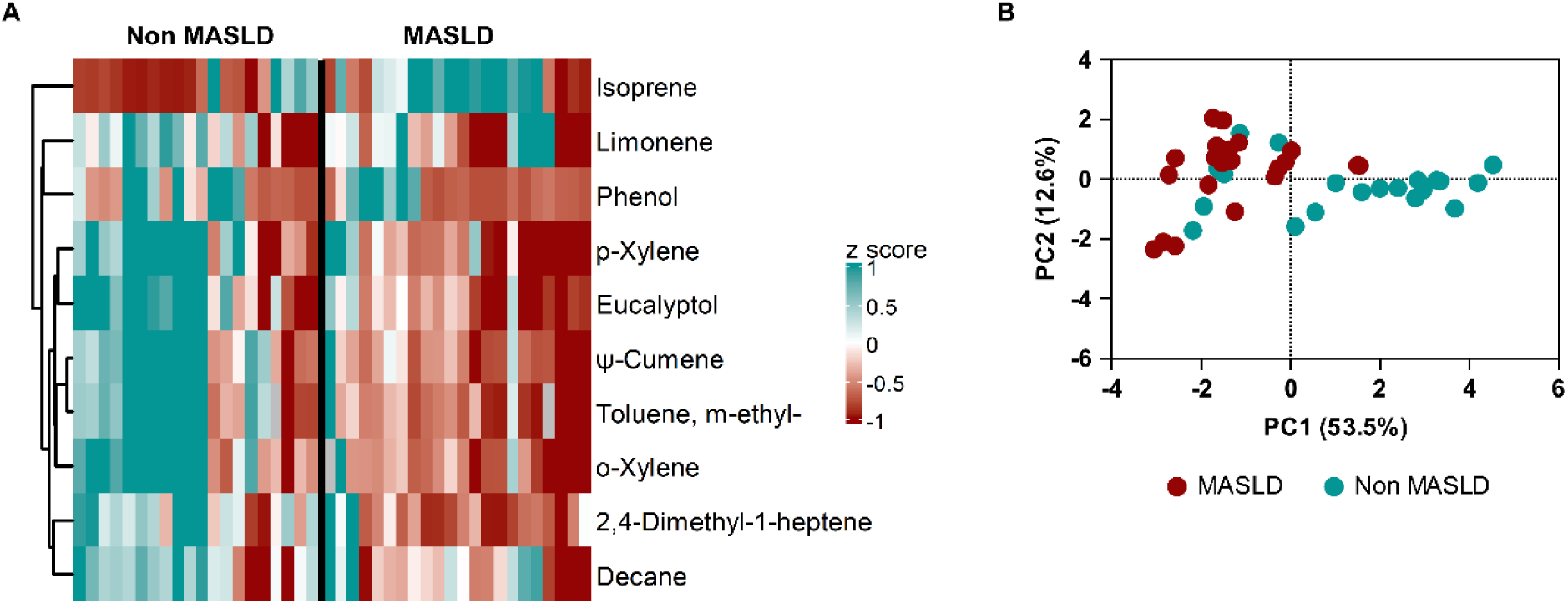
Ten breath VOCs discriminate MASLD children from Non MASLD. **A)** *z* score abundances of ten discriminant volatiles in the breath of children with and without MASLD. The color code (teal to dark red) displays z score: teal, high abundance; red, low abundance**. B)** Principal components analysis to visualize the overall difference in breath composition in children with and without MASLD.

Using the 10 discriminatory VOC biomarkers, a Random Forest classifier was built. The resulting model yielded a sensitivity of 73% and a specificity of 65%, AUC 0.84 (**Figure 2A and 2B**). Importantly, a single biomarker, 2,4-dimethyl-1-heptene, distinguished MASLD from non MASLD with an AUC of 0.81, sensitivity of 85%, and a specificity of 77% (**Figure 2C and 2D**).

**Figure 2:**
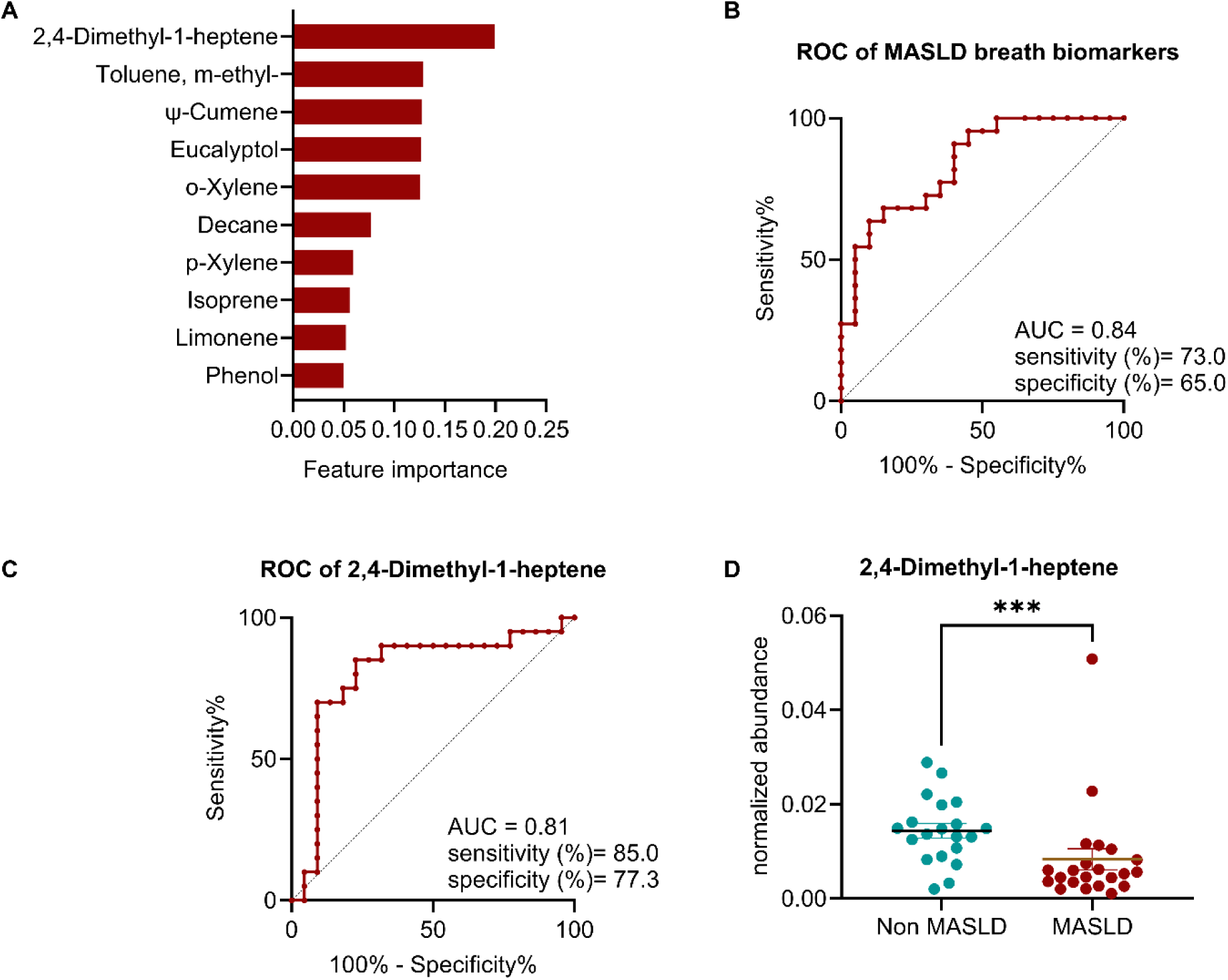
Predictive value of breath VOCs for diagnosis of MASLD. **A)** Feature importance of the ten discriminant volatiles determined using Random Forest Classifier with Gini importance. **B)** Receiver operating characteristic (ROC) curve model using 10 MASLD breath VOCs and **C)** using 2,4-dimethyl-1-heptene alone to predict MASLD. **D)** Normalized abundances of 2,4-dimethyl-1-heptane in the breath of children with and without MASLD.

### Hierarchical Clustering of MASLD Patients Based on VOC Profiles

Substantial heterogeneity was noted in the breath volatilome of subjects with MASLD. We hypothesized that this heterogeneity was due to biological differences between subjects. Using all discriminant VOCs, we performed hierarchical clustering on subjects with MASLD, identifying four clusters (**Figure 3A**). Cluster 1 contained three subjects, cluster 2 contained ten subjects, cluster 3 had a single subject, and the remaining subjects formed cluster 4. Subjects in cluster 1 had the lowest VOC abundance. Cluster 2 subjects were characterized by elevated breath isoprene levels, whereas cluster 4 patients demonstrated elevated levels of breath phenol (**Figures 3A and 3B**). The single patient in cluster 3 displayed consistently higher levels of most aromatic hydrocarbons. We next interrogated whether cluster assignments were associated with clinical or laboratory results. We found that patients in cluster 1 had the lowest mean ALT, AST, and GGT values and the highest mean ALP levels (**Supplementary Figure 1**). Patients in cluster 2 had the highest mean ALT and AST levels. Patients in cluster 4 had significantly higher total protein levels. Correlation studies for cluster 2 showed that there was a significant and strong positive correlation between the most predictive VOC—2,4-dimethyl-1-heptene—and GGT levels (r= 0.73, p=0.031), in addition to a strong positive correlation between phenol and total bilirubin (r= 0.75, p=0.015) (**Figure 4A**). In cluster 4, strong correlations were also identified between glucose and phenol (r= 1.00, p=0.016), and between BMI z-score and 2,4-dimethyl-1-heptene (r= 0.91, p=0.004), while o-xylene was negative correlated to glucose (r=-0.9, p=0.08) (**Figures 4B)**. Correlations for other clusters (cluster 1 & 3) were not performed, due to the limited number of samples and/or missing laboratory results.

**Figure 3:**
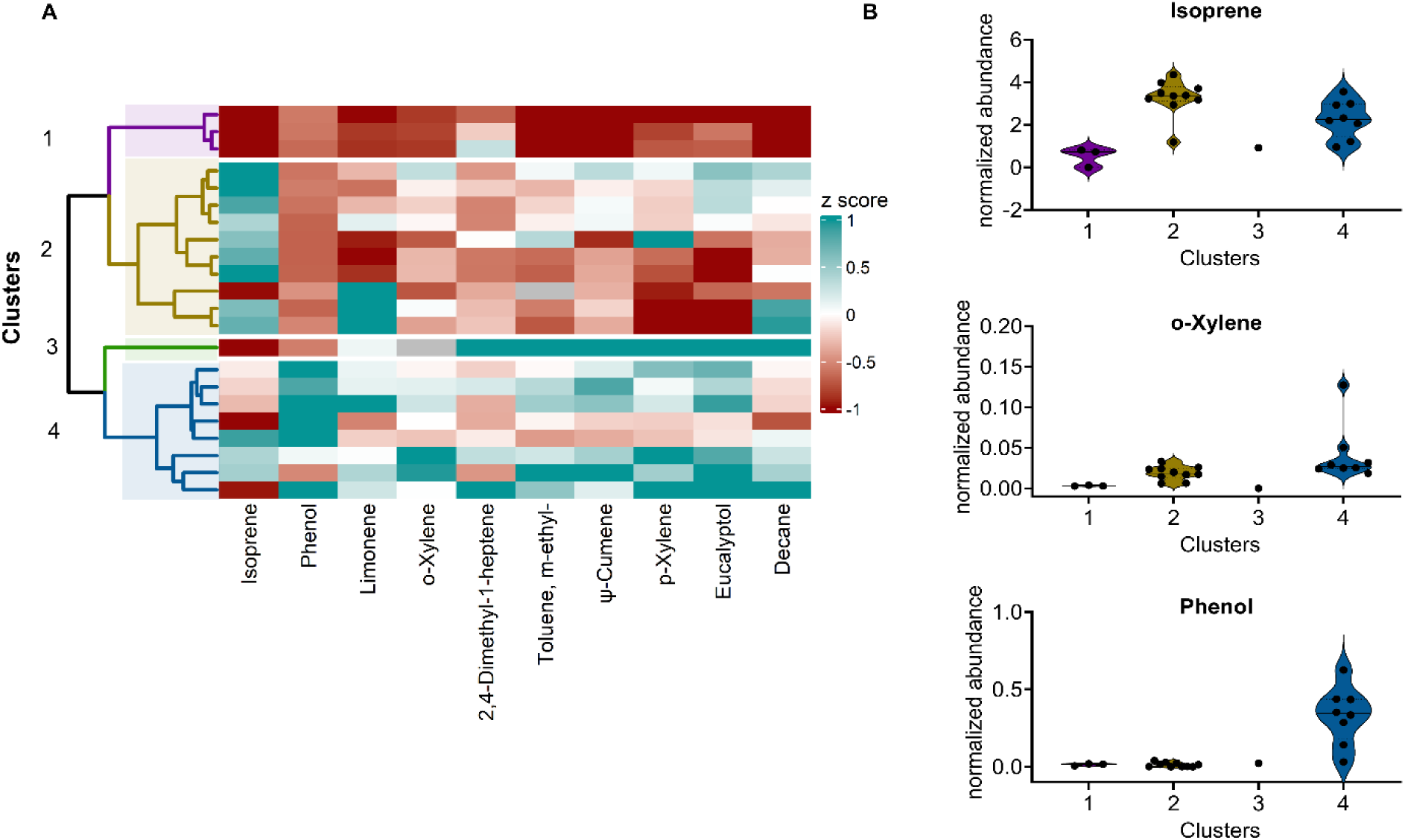
Hierarchical clustering identifies breath VOC phenotypes in patients with MASLD. **A**) heatmap visualizes the relative abundance of each breath VOC (columns) for each subject with MASLD (rows). The color code (teal to dark red) displays z score: teal, high abundance; red, low abundance. The dendrogram shows hierarchical clustering for all subjects with MASLD, based on the Euclidean distance and Ward’s cluster agglomeration method. **B**) Distribution of specific MASLD VOCs, by cluster.

**Figure 4:**
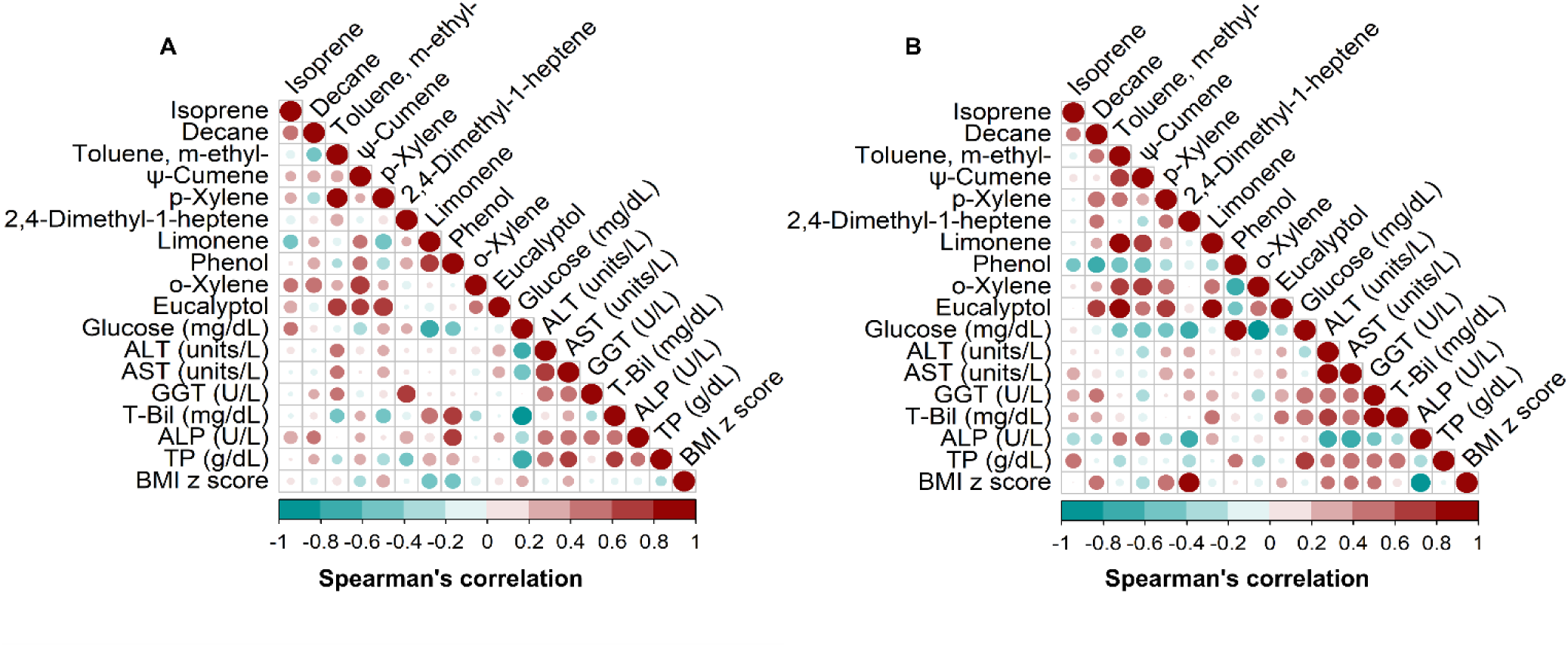
Correlations between breath MASLD VOCs and clinical features for cluster 2 (A) and cluster 4 (B). ALT= Alanine aminotransferase. AST= Aspartate aminotransferase. GGT = gamma-glutamyltransferase. T-Bil = Total Bilirubin. ALP = Alkaline phosphatase. TP = Total Protein.

### Histological Features Correlate with MASLD-Associated VOCs

Among participants who underwent liver biopsy, histological analysis revealed that 6 out of 10 subjects had no evidence of fibrosis, while 8 subjects had a NAFLD activity score (NAS) between 3 and 4, values typically classified as borderline MASH. Although NAS scoring was evaluated (**Supplementary Table 1**, **Supplementary Figure 2**).

To explore whether breath VOC concentrations were linked to histological disease severity, we performed Spearman’s correlation analyses between individual VOCs and both fibrosis stage and NAFLD Activity Score (NAS). Correlation strength was quantified using the correlation coefficient (r; range −1 to 1), with values closer to zero indicating weaker linear associations, and statistical significance was assessed using the corresponding p value to determine whether the observed correlation differed from zero (**Figures 5A and 5B**). Among the detected compounds, 2,4-dimethyl-1-heptene was strongly positively correlated with fibrosis stage (r = 0.72, p = 0.026). Phenol and eucalyptol showed a moderate-to-strong correlation with NAS (r = 0.69, p = 0.03 and r = 0.64, p = 0.05 respectively) (**Supplementary Figure 2**). These associations suggest that higher levels of 2,4-dimethyl-1-heptene, phenol, and eucalyptol in breath may reflect greater histological severity, particularly with respect to overall disease activity, and warrants further investigation as a candidate marker of advanced disease.

**Figure 5:**
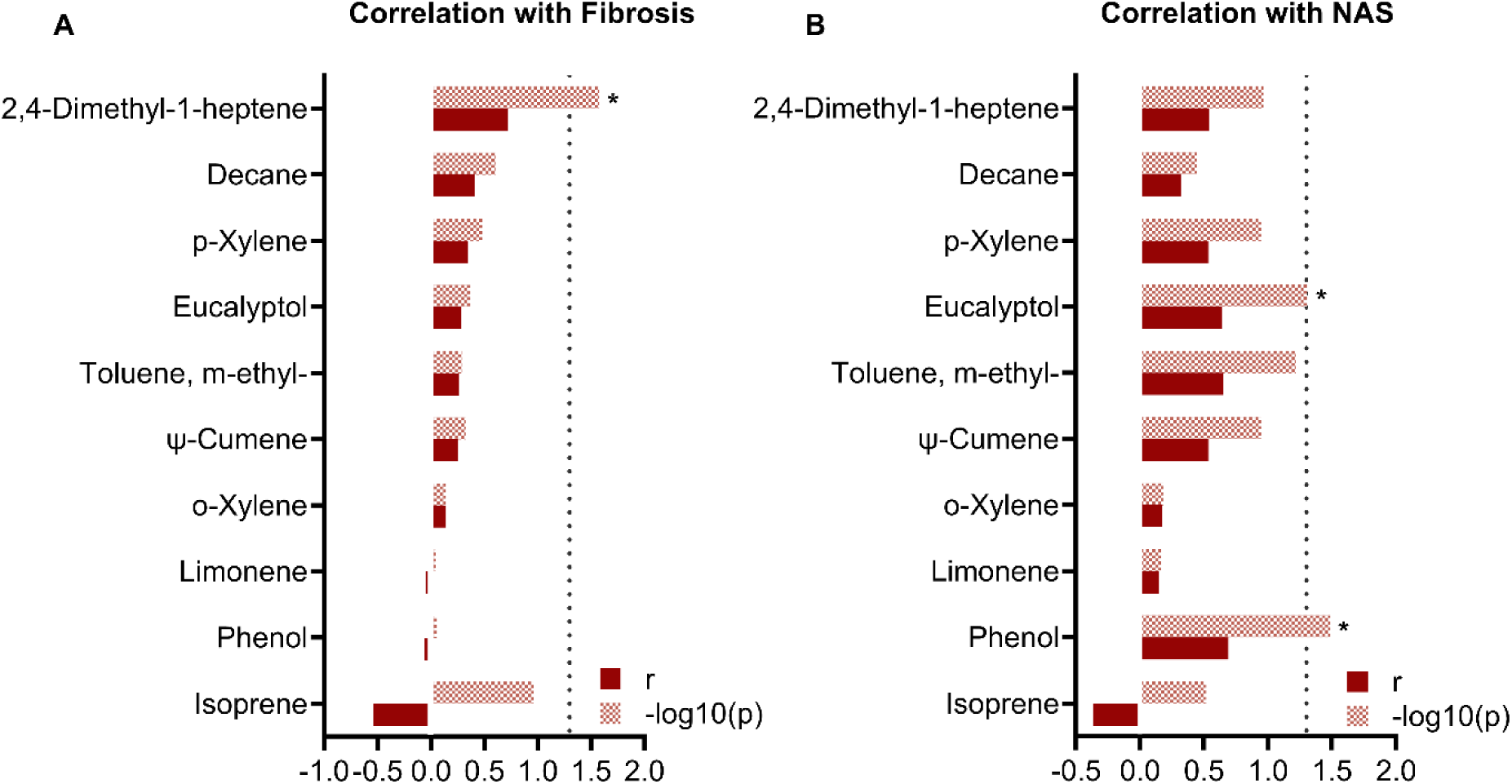
Association between disease severity and breath metabolite levels. Correlation plots for fibrosis **(A)** and NAFLD activity score (NAS) **(B)** for all features from exhaled breath (dotted line indicates -log10(p=0.05), asterisks denote correlations with p<0.05).

Interestingly, while non MASLD exhibited higher relative levels of some of these VOCs compared to MASLD patients, within MASLD individuals, elevated VOC levels correlated with greater disease severity. This pattern highlights the complexity of interpreting VOCs: they may act as markers of both metabolic status and disease progression, depending on the clinical context.

### o/p-Xylene is a reproducible MASLD indicator

To evaluate the reproducibility of our findings, we took advantage of samples previously acquired during an independent study performed at St. Louis Children’s Hospital (SLCH). All children in this validation cohort had overweight or obesity, with a BMI of at least 85% for age (**Supplementary Table 2**). There were no significant differences in age, sex and race between MASLD patients and non MASLD. Consistent with disease pathology, AST and ALT levels were significantly elevated in MASLD patients compared to controls.

Of ten VOCs identified in the CHOP MASLD cohort, four of these—isoprene, limonene, o/p-xylene (ortho- and para-structural isomers of xylene were not chromatographically resolved), and phenol—were detected in breath samples from the SLCH cohort (**Supplementary Figure 3**). Other MASLD VOCs were not detected, which was not unexpected due to differences in instrument sensitivity. Among these four VOCs, only o/p-xylene demonstrated a statistically significant difference between patients under evaluation for MASLD and non MASLD, consistent with findings from the primary cohort (**Figures 6A and 6C, Supplementary Figure 4**). In the primary CHOP cohort, the combined abundance of p-xylene and o-xylene yielded an AUC of 0.82, with a sensitivity of 95% and specificity of 54.5% (**Figures 6B)**. When applied to the validation dataset (at SLCH), prediction of MASLD diagnosis achieved an AUC of 0.78, with sensitivity and specificity for this compound of 93% and 53% respectively (**Figures 6D, Supplementary Figure 4**). These results suggest that o/p-xylene is a reproducible breath biomarker of MASLD across independent cohorts.

**Figure 6:**
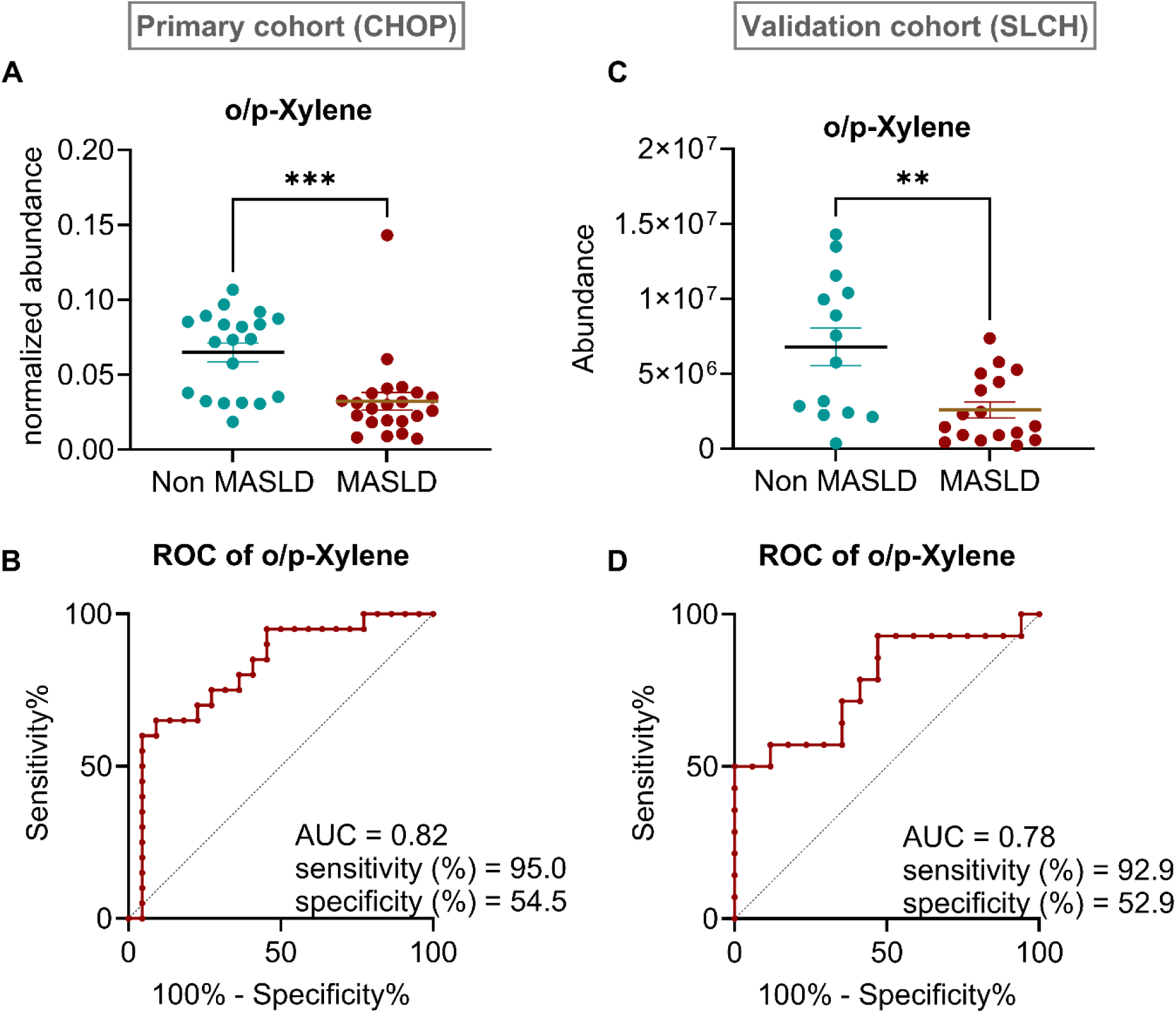
Breath abundance of xylene is reproducibly reduced in the breath of children with MASLD. Normalized breath abundances of cumulative o/p-xylene and receiver operating characteristic (ROC) curve model to predict MASLD in a primary discovery cohort **(A-B)** and independent validation cohort **(C-D)**.

## DISCUSSION

Even in the presence of cardiometabolic risk factors, elevated ALT alone is insufficient for the diagnosis of pediatric MASLD. Current diagnosis pathways require confirmation of hepatic steatosis via imaging or liver biopsy, in combination with at least one cardiometabolic risk factor. Liver biopsy, historically thought to be the gold standard, is no longer standard for MASLD diagnosis, as its use is limited due to its invasive nature, procedural risk, cost, and susceptibility to sampling error. Noninvasive serum biomarkers such as the Fibrosis-4 Index (FIB-4) are considered the most cost-effective first-line screening tools for MASLD in adults. However, FIB-4 demonstrates poor diagnostic performance in pediatric populations and is not a validated marker for children. Serum ALT remains the most widely accepted and validated screening biomarker in pediatric MASLD because of its accessibility, with reported sensitivity and specificity of approximately 80–85% with an AUC of 0.66–0.68. However, ALT has important limitations, as up to 25–30% of patients with biopsy-proven MASH or hepatic steatosis may have normal ALT levels. Imaging modalities, while more acceptable, can be confounded by factors such as inflammation, fibrosis, and obesity ^1^. Thus, there is an urgent need for additional noninvasive biomarkers to screen and stratify children with MASLD.

Breath-based VOC testing represents a promising, noninvasive approach that could be deployed as a portable, rapid, point-of-care screening tool in children who meet screening guidelines for MASLD, perhaps even at a routine well child appointment. Children with an abnormal breath signature and/or abnormal ALT would then be triaged to targeted imaging studies, reserving liver biopsy only as needed for diagnostic confirmation or assessment of disease severity.

Numerous studies have investigated breath profiles in adults with liver diseases including MASH, hepatic encephalopathy, hepatocellular carcinoma, or cirrhosis, typically in comparison to healthy controls, using GC-MS or PTR-MS technologies ^18–22^. Breath VOCs associated with liver disease have included terpenes, ketones, aromatic compounds, and sulfur-containing compounds. Diagnostic performance has generally been high, with sensitivities ranging from 83% to 100% and specificities from 70% to 100%. The only studies reported on pediatric population differentiate cirrhotic children and NAFLD from healthy children using the SIFT-MS ^10,11^ and focused on a small set of preselected VOCs.

To our knowledge, this is the first study to apply untargeted GCxGC-MS analysis to elucidate and validate features that discriminate pediatric MASLD. Several of the biomarkers found in our pediatric cohort have been previously reported in adult breath studies of cirrhosis or hepatic encephalopathy, although the direction of change often differed from our findings, likely reflecting age-related differences in hepatic metabolism.

Unsupervised clustering analysis identified four distinct subgroups of children with MASLD, characterized by differing volatile phenotypes and associated clinical features. One subgroup was characterized by globally lower abundances of discriminatory VOCs alongside lower serum liver transaminases, suggesting a milder or less active disease phenotype —potentially representing patients at lower risk who may not require aggressive intervention but still warrant long-term monitoring. In contrast, another cluster exhibited elevated breath isoprene levels in conjunction with higher ALT and AST, suggesting a phenotype associated with increased hepatic injury or inflammatory activity. Within this group, correlations between specific VOCs (e.g., 2,4-dimethyl-1-heptene and phenol) and serum markers such as GGT and total bilirubin further support a potential link between breath VOCs and underlying liver dysfunction. Additional clusters revealed associations between elevated breath phenol levels and higher total serum protein, as well as positive correlations between phenol and glucose and between BMI and 2,4-dimethyl-1-heptene, highlighting the potential of breath profiling to capture metabolic heterogeneity within pediatric MASLD. Collectively, these findings suggest that VOC profiling not only distinguishes MASLD from non MASLD children, but also further delineates clinical meaningful subgroups with MASLD. This capacity for sub-stratification suggests that VOC analysis captures disease heterogeneity beyond current diagnostic tools. Larger, longitudinal studies will be required to validate these clusters and define their prognostic significance.

Using the same standardized breath collection protocol, we evaluated whether key biomarkers identified in the discovery cohort were reproducible in an independent pediatric population at SLCH. o/p-Xylene demonstrated reproducible, directionally consistent changes across cohorts, with significantly lower levels observed in children with MASLD compared with non MASLD controls. Xylenes are environmentally ubiquitous aromatic hydrocarbons, primarily encountered through inhalation, and are highly lipophilic, readily partitioning into adipose tissue ^23^. This physicochemical property provides a plausible biological explanation for their altered breath levels in MASLD. One hypothesis is that increased liver adiposity in MASLD serves as a storage reservoir for xylenes, reducing their availability for pulmonary exhalation. Alternatively, MASLD-associated gut dysbiosis may modulate xylene bioavailability through enhanced microbial degradation or transformation of aromatic compounds, as suggested by emerging microbiome studies ^24–26^. These findings indicate that breath concentrations of o/p-xylene may reflect a combination of environmental exposure, host metabolic state, and microbiome interactions, highlighting its potential utility as a reproducible, biologically informative breath biomarker for pediatric MASLD.

## Strengths and Limitations

This study has several key strengths. The CHOP cohort was rigorously phenotyped, with determination of MASLD through imaging, laboratory blood work, and liver biopsy results when available. The comparator group consisted of at-risk children with elevated BMI, allowing discrimination of MASLD-specific breath signatures from obesity-related effects. Importantly, key findings, including reduced o/p-xylene in MASLD group, were reproducible across two independent pediatric cohorts at CHOP and SLCH, supporting the robustness of these findings.

There are several limitations to our study. The two study sites employed different mass spectrometry instruments with varying sensitivities, which is expected to influence the detection of low abundance VOCs, which could include those with the highest predictive value. A potential confounder is that the CHOP cohort was not fully matched for sex, of concern given the higher risk of MASLD progression in boys; however, no sex differences were observed in the validation cohort, partially mitigating this issue. Both cohorts had relatively small sample sizes, and demographic differences existed between cases and controls in the primary CHOP cohort. Additionally, the SLCH validation cohort was less extensively phenotyped than the discovery cohort, limiting direct clinical comparisons. Despite these limitations, the reproducibility of key biomarkers across independent populations reinforces the potential translational relevance of breath VOC analysis for pediatric MASLD.

## CONCLUSIONS

Pediatric MASLD remains underdiagnosed due to the lack of accurate, noninvasive diagnostic tools suitable for early disease detection and longitudinal monitoring. Breath-based VOC analysis has the potential to address this critical gap by offering a rapid, child-friendly approach that could be implemented as a point-of-care screening test to identify at-risk children and guide downstream diagnostic evaluation. In this study, we demonstrated that children with MASLD exhibit a distinct breath VOC signature, with 2,4-dimethyl-1-heptene showing strong diagnostic performance in the discovery cohort (85% sensitivity, 77% specificity, AUC 0.81). Exploratory clustering revealed heterogeneity in breath profiles from MASLD patients, suggesting opportunities for disease stratification beyond binary diagnosis. Importantly, key biomarkers, including o/p-xylene, were reproducibly validated in an independent pediatric cohort, supporting their robustness and biological relevance. Together, these findings establish breath VOC profiling as a promising, noninvasive strategy for pediatric MASLD screening and risk stratification, warranting validation in larger, longitudinal studies.

## Data Availability

All data produced in the present study are available upon reasonable request to the authors

## ACKNOWLEDGMENT

We acknowledge funding from the OMICS Maximizing Grant, Children’s Hospital of Philadelphia. Support was also received through the National Institutes of Health (NIH) National Institute of Allergy and Infectious Diseases (R21 AI154370 to A.R.O.J.), and National Institute of Child Health and Human Development (R01 HD109963 A.R.O.J.)

## Notes

### Competing Interest Statement

The authors have declared no competing interest.

### Author Declarations

Childrens Hospital of Philadelphia Human Research Ethics Committee (IRB 21 019125) gave approval for this work. St. Louis Childrens Hospital Human Research Ethics Committee (IRB 201709030) gave approval for this work.

