## Supplemental Files for "Breath volatile profiling reveals a diagnostic signature of MASLD in children"

*Corresponding author: Audrey R. Odom John

**Supplementary Figure 1:** Distribution of clinical features by cluster

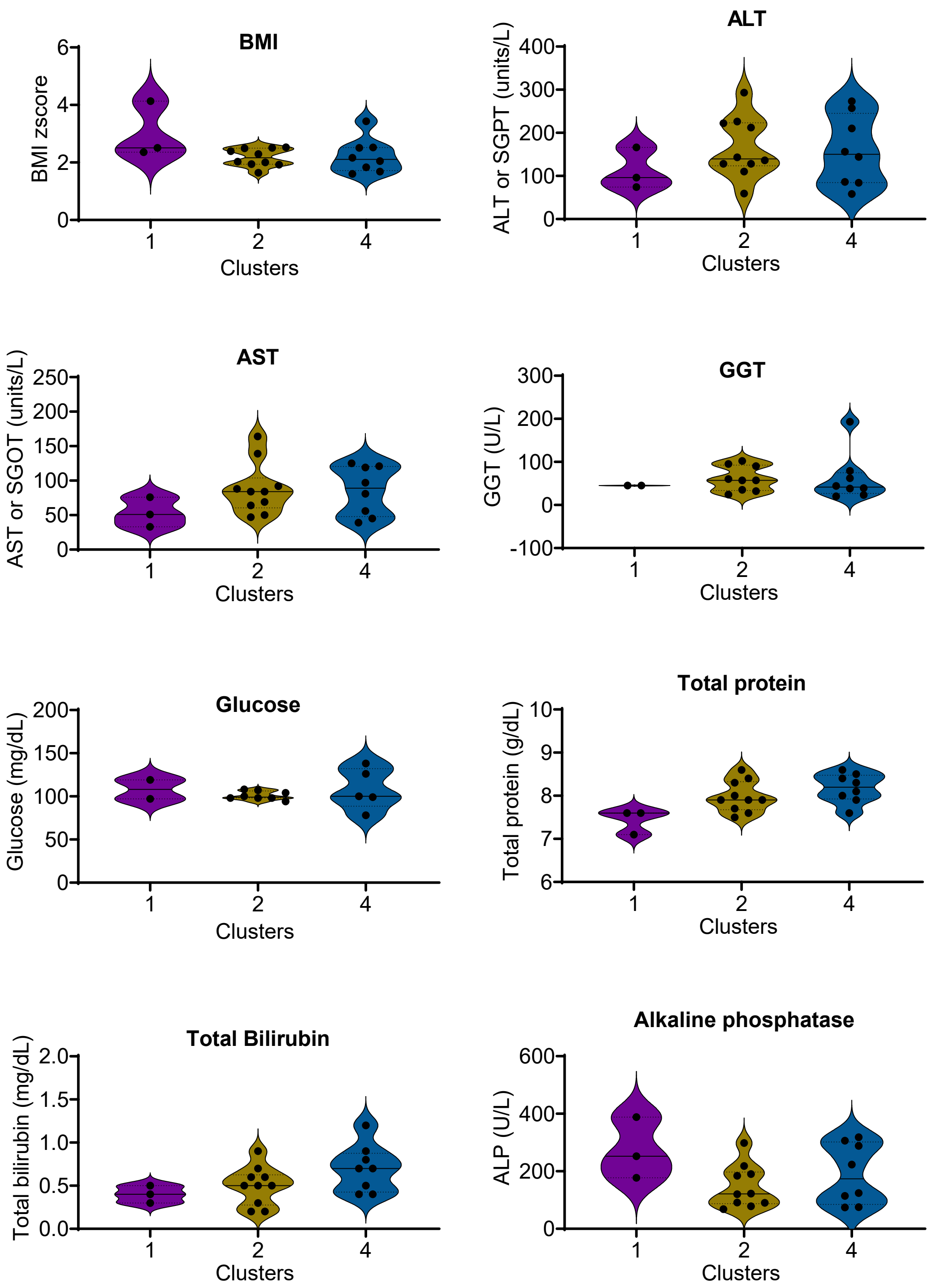

**Supplementary Figure 2: A-B)** NAFLD activity score (NAS) and fibrosis scores for biopsy samples from subjects with MASLD. **C)** Normalized abundance of predictive VOC in subjects with and without MASLD.

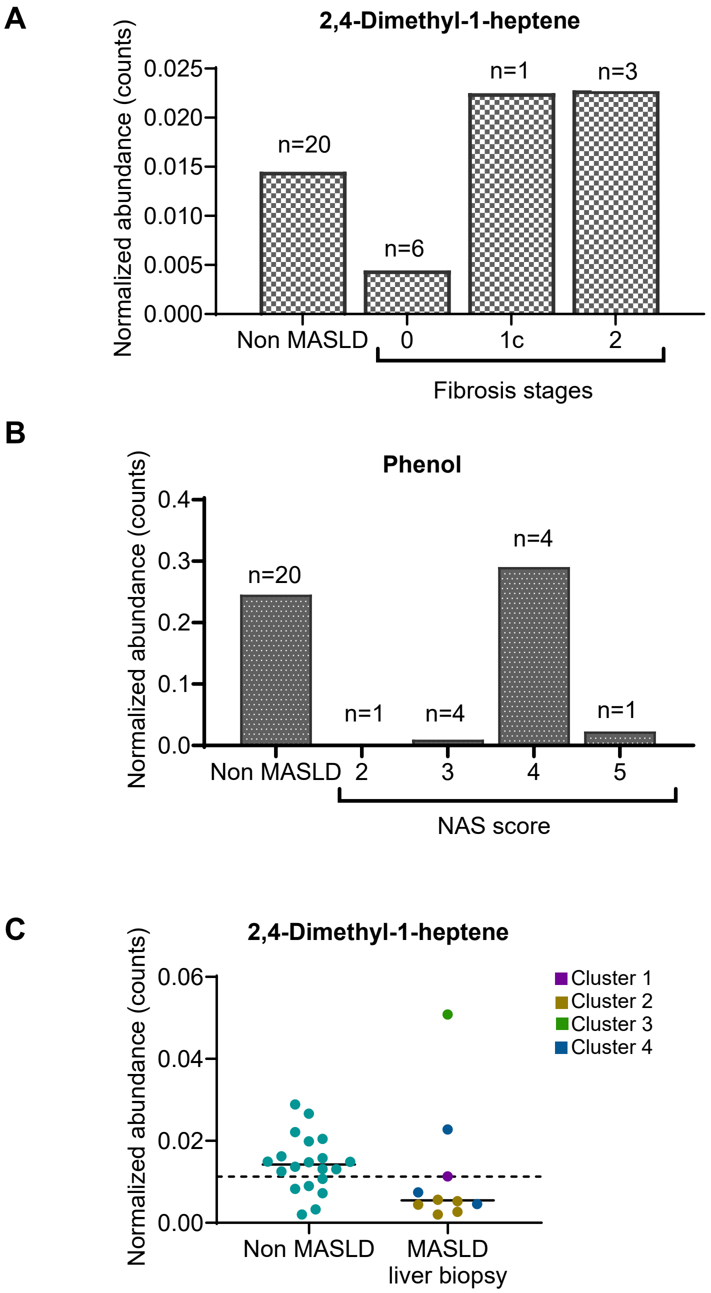

**Supplementary Figure 3:** MASLD-associated VOCs detected in validation cohort (SLCH)

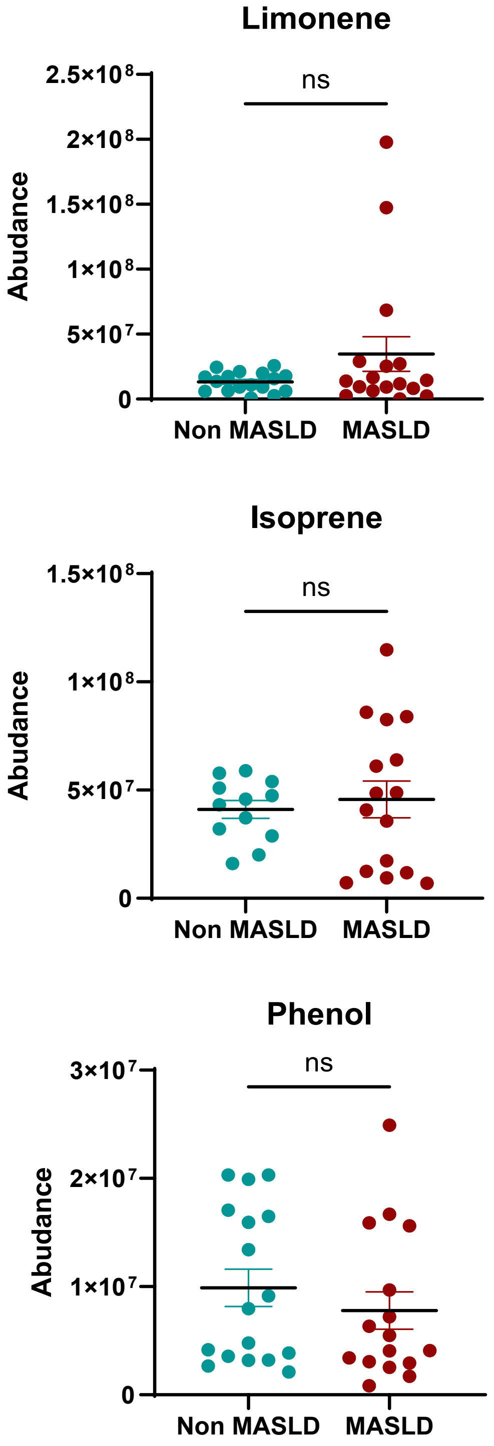

**Supplementary Figure 4: A-B)** Scatter plots showing normalized breath abudance of o-xylene and p-xylene (primary cohort, CHOP) and **C-D**) receiver operating characteristic (ROC) curve using o-xylene and p-xylene to predict MASLD.

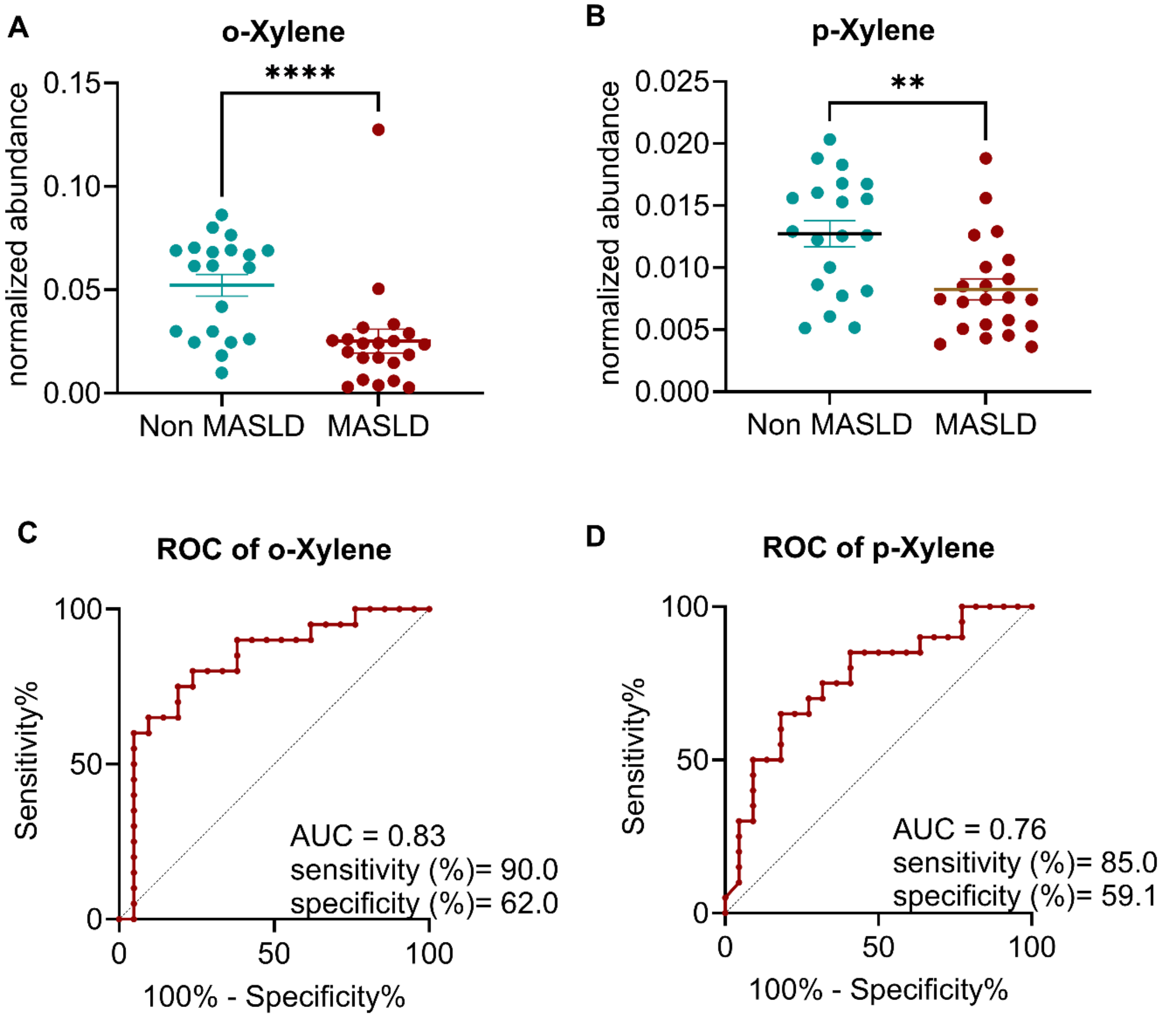

**Supplementary Table 1.** Histological charateristics of biopsy-proven MASLD.

| Liver Histological Features | | |
|---|---|---|
| MASLD subjects (n=10) | NAFLD Activity Score (0-8) | Fibrosis stage (0-4) |
| Subject 1 | 3 | 1c |
| Subject 2 | 3 | 0 |
| Subject 3 | 4 | 2 |
| Subject 4 | 5 | 1c |
| Subject 5 | 4 | 0 |
| Subject 6 | 2 | 0 |
| Subject 7 | 3 | 0 |
| Subject 8 | 3 | 1c |
| Subject 9 | 4 | 0 |
| Subject 10 | 4 | 0 |

**Supplementary Table 2.** **Characterization of validation study population by MASLD status (SLCH cohort).**

| Variable | Non MASLD,<br>N = 18[1] | Under evaluation for MASLD,<br>N = 18[1] | p-value[2] |
|---|---|---|---|
| **Clinical Features** | | | |
| Age | 11.5 (10.00, 14.00) | 14 (11.25, 16.75) | 0.074 |
| Sex | | | 0.2 |
| Female | 13 (72%) | 9 (50%) | |
| Male | 5 (28%) | 9 (50%) | |
| Race | | | 0.5 |
| Black or African American | 4 (22%) | 1 (5.6%) | |
| Other | 1 (5.6%) | 1 (5.6%) | |
| White American / Caucasian | 13 (72%) | 16 (89%) | |
| BMI z score | 2.65 (2.05, 3.09) | 2.57 (1.97, 3.48) | 0.9 |
| BMI | 36 (28, 40) | 36 (29, 41) | 0.7 |
| Glucose (mg/dL)<br>n available | 95 (86, 102)<br>16/18 | 88 (85, 96)<br>18/18 | 0.6 |
| ALT (units/L)<br>n available | 23 (19, 28)<br>15/18 | 94 (76, 118)<br>18/18 | <0.001 |
| AST (units/L)<br>n available | 20 (17, 27)<br>14/18 | 55 (45, 70)<br>18/18 | <0.001 |

[1]Median (IQR); n (%). [2]Wilcoxon rank sum test; Pearson's Chi-squared test; Fisher's exact test. ALT= Alanine aminotransferase. AST= Aspartate aminotransferase.
